# Impact of Acute In-Hospital Post-Burn Pruritus on Quality of Life: A Cross-Sectional Study

**DOI:** 10.1101/2025.04.20.25326122

**Authors:** Mohammad Tolouei, Sanaz Masoumi, Mehrnaz Kooshanfar, Ehsan Kazemnezhad Leyli, Ramyar Farzan

## Abstract

**Background:** Post-burn itching is a common and distressing complication after thermal injury that can severely diminish patients’ well-being. Despite its prevalence, the acute effects of itch during hospitalization and how it relates to specific burn characteristics remain insufficiently explored, particularly in non-Western populations.

**Methods:** We conducted a cross-sectional study in 2021 at Velayat Hospital in Rasht, Iran, enrolling 268 adults who had been admitted for at least 48 hours following a burn injury. Through patient interviews and medical-record review, we gathered information on demographics; burn severity—measured by the percentage of total body surface area affected, depth of tissue damage, and anatomical location of burns; and itch parameters—measured by patient-rated intensity on a 0–10 scale, duration in days, and areas of the body affected. To assess long-term well-being, we used the ItchyQoL questionnaire, which quantifies the impact of itching on physical, emotional, and functional aspects of quality of life. We applied parametric, non-parametric comparisons and multivariable regression models to examine how itch characteristics and burn severity influenced immediate recovery and subsequent quality of life.

**Results:** Patients who experienced itching had larger areas of skin damage, deeper tissue injuries, and longer hospital stays than those without itching. Higher itch intensity and longer itch duration both correlated with poorer scores on the ItchyQoL, indicating a greater negative impact on quality of life. In multivariable analysis, itch intensity emerged as the strongest predictor of reduced well-being, followed by duration of itching, depth of the burn injury, and percentage of body surface area burned. Moreover, the anatomical site of the burn influenced both the severity of itching and the length of hospitalization.

**Conclusions:** The intensity and persistence of itching during the acute hospitalization period play a critical role in patients’ recovery trajectories and their long-term quality of life after a burn injury. Early recognition and aggressive management of severe or prolonged itching may help to shorten hospital stays, enhance recovery, and improve overall well-being. The complete dataset, analysis code, and detailed findings are publicly accessible in our [GitHub](https://github.com/ramyarfarzan/Impact-of-Acute-In-Hospital-Post-Burn-Pruritus-on-Quality-of-Life-A-Cross-Sectional-Study.git) project repository.

## 1 Introduction

Burn injuries present a significant global health issue, and postburn pruritus has emerged as one of the most enduring and troublesome complications. Up to 93% of burn patients report experiencing acute itching during the early recovery phase, while many continue to suffer from chronic symptoms that interfere with sleep, daily activities, and psychological well-being [10, 16]. Current evidence suggests that both peripheral factors, such as the depth of burns and the need for skin grafting, and psychological factors, including post-traumatic stress, are involved in modulating the severity of itch [3].

Although many studies have focused on chronic itch outcomes and evaluated specific treatment protocols [24, 5, 20, 17, 4, 26], limited attention has been given to how itching during the acute phase of hospitalization is related to burn characteristics and its influence on long-term quality of life. This gap is important because early experiences of itch can shape both the course of physical recovery and the process of psychological adjustment after injury [12, 14]. Moreover, only a few studies have applied burn-specific questionnaires that can capture the unique and complex effects of itch [8, 25].

The development of validated tools, such as the ItchyQoL questionnaire [23] (adapted and validated in Persian), offers a promising way to measure the subjective burden of itch across physical, emotional, and functional aspects. Recent studies highlight the importance of these detailed instruments in uncovering complex relationships between physiological factors (e.g., TBSA, burn depth), demographic factors (e.g., age, gender), and itch-specific characteristics (e.g., severity, duration). These insights can help improve the predictive accuracy of recovery models [21, 22].

The impact of post-burn pruritus on quality of life is also well documented. Chronic itch has been linked to significant disturbances in sleep, heightened psychological distress, and diminished physical functioning [3]. Yet, most of these investigations rely on generic quality-of-life assessments that often overlook burn-specific issues, such as the discomfort from scarring or the social stigma associated with burn injuries. Recent advancements, including burn-specific instruments like the ItchyQuant and ItchyQoL questionnaires, offer a more tailored evaluation of these effects, though their application has predominantly been limited to the chronic phase of recovery [7, 23].

Moreover, there exists a clear regional bias in the literature that has been documented. Most of the current results are based on research conducted in Western populations, while evidence from other areas, such as Iran, is still limited [1]. The geographical discrepancy raises questions about the generalizability of the results to different cultural and healthcare settings and underscores the importance of conducting context-specific studies.

To address these gaps, the present study aims to explore the characteristics of pruritus during the acute phase of hospitalization. By investigating the relationship between early itch severity and burn-related factors, such as total body surface area, burn depth, and anatomical location, and by applying a burn-specific quality-of-life measure, this study seeks to expand current understanding and support the development of targeted interventions that can improve short-term patient outcomes.

## 2 Material and Method

### 2.1 Study Design

This study is a cross-sectional research conducted to investigate the relationship between burn-related itching during hospitalization, burn characteristics, and its impact on quality of life (QoL) among burn patients. Data were collected from patient records during their hospital stay, focusing on individuals admitted for at least 48 hours. This design provided an opportunity to assess itching and its effects on QoL during the acute phase of burn recovery, in line with previous studies on post-burn pruritus and its influence on patient outcomes [18, 6, 19].

### 2.2 Participants

The study included 268 adult burn patients admitted to Velayat Hospital in Rasht, Iran, in 2021. The inclusion and exclusion criteria—adapted from previous research works [13, 2, 9, 11]—were designed to ensure that the observed outcomes, particularly regarding itch characteristics and post-burn quality of life, were attributed primarily to the burn injury rather than to other confounding factors.

Participants were selected based on the following inclusion criteria: (1) provision of written informed consent; (2) hospitalization for at least 48 hours; (3) age between 18 and 64 years; and (4) availability of complete data on itch reporting, itch severity, itch duration, and quality of life. The exclusion criteria were: (1) presence of dermatological conditions such as dermatitis, psoriasis, or acne; (2) diagnosis of acute psychosis; (3) burns caused by self-immolation; (4) use of medications that could influence itching, including morphine, topical steroids, topical antihistamines, or other topical agents; (5) use of immunosuppressive treatments such as systemic steroids or chemotherapy; and (6) alcohol or drug abuse. Additionally, the sample was adjusted to ensure equal gender representation (50% male, 50% female) to reduce potential bias.

### 2.3 Data Collection Procedure

Data were collected at Velayat Hospital using structured patient interviews and systematic reviews of medical records. All interviews were conducted by trained research staff to ensure consistency. To promote accurate symptom reporting, interviews were carried out at a single, standardized time point during hospitalization, typically after the initial 48 hours of admission, allowing time for patient stabilization. Each participant was assigned a unique identifier to maintain confidentiality and support organized data analysis.

The following variables were recorded for each patient: (1) **Age**, recorded in years at the time of admission, based on hospital records; (2) **Gender**, self-identified by the patient as Male or Female and verified through hospital records; (3) **Total Body Surface Area (TBSA) Burned**, assessed by specialized burn clinicians using standardized methods such as the Lund-Browder chart [15], expressed as a percentage of the body surface area affected (ranging from 1% to 50%); (4) **Burn Depth**, classified as Partial-thickness or Full-thickness based on clinical examination by burn specialists; (5) **Burn Location**, documented as the anatomical regions affected by the burn, such as Upper Limbs, Lower Limbs, Trunk, or combinations thereof, determined through clinical assessment; (6) **Accident Place**, the setting where the burn incident occurred (e.g., Home, Work), as reported by the patient or family members during interviews; (7) **Burn Cause**, determined through clinical history and physical examination, categorized as Thermal, Electrical, or Chemical; (8) **Comorbidities**, pre-existing conditions recorded from medical records, including Diabetes, Hypertension, Cardiovascular, Renal, or None, to account for potential confounding factors.

Additional variables related to itch and quality of life were also recorded: (9) **Itch Report**, a binary indicator (Yes or No) of whether the patient experienced itching during hospitalization, based on patient self-reports during interviews; (10) **Itch Severity**, quantified using the Itch-Man Scale, a validated 0–10 Numerical Rating Scale (NRS), where 0 represents no itch and 10 represents the worst possible itch, with patients who did not experience itching assigned a score of 0; (11) **Itch Duration**, measured as the average number of hours per day the patient experienced itching (ranging from 0 to 24 hours), based on patient self-reports, with patients reporting no itching assigned a duration of 0; (12) **Itch Location**, the anatomical regions affected by itching, such as Trunk, Upper Limbs, Lower Limbs or combinations, often corresponding to burn sites, as reported by the patient; (13) **Quality of Life (QoL) Score**, evaluated using the ItchyQoL questionnaire, a validated 26-item instrument assessing the impact of pruritus on quality of life across three domains: symptoms, functioning, and emotions, with each question scored on a 5-point Likert scale (1 = never, 5 = always), yielding a total score ranging from 26 to 130, where higher scores indicate worse quality of life due to itching; (14) **Hospitalization Duration**, the total number of days from admission to discharge, ranging from 2 to 19 days in this study sample (though the acceptable range per study design was 2 to 30 days), extracted from hospital records. All data were de-identified to protect patient confidentiality, and the collection process was designed to ensure accuracy and consistency across all participants. A summary of the variables, their descriptions, and their ranges of values is provided in Table 1.

**Table 1:** List of Variables. Each variable is accompanied by a brief description and its range of values in the dataset.

| Variable | Description | Range of Values |
| --- | --- | --- |
| Age | Patient’s age in years at the time of admission | 18–64 |
| Gender | Patient’s self-identified gender, verified via hospital records | Male, Female |
| TBSA_Burned | Percentage of total body surface area affected by burns, assessed using Rule of Nines or Lund-Browder chart | 1–50 |
| Burn_Depth | Clinical classification of burn severity based on depth | Full-thickness, Partial-thickness |
| Burn_Location | Anatomical regions affected by the burn, determined clinically | Upper Limbs, Lower Limbs, Trunk, Combinations |
| Accident_Place | Setting where the burn incident occurred, reported by patient or family | Work, Home, Other |
| Burn_Cause | Cause of the burn, determined through clinical history and examination | Thermal, Electrical, Chemical |
| Comorbidities | Pre-existing conditions recorded from medical records | Cardiovascular, Diabetes, Renal, Hypertension, None |
| Itch_Report | Binary indicator of whether the patient experienced itching during hospitalization | Yes, No |
| Itch_Severity | Severity of itching measured using the Itch-Man Scale (0 = no itch, 10 = worst possible itch) | 0–10 |
| Itch_Duration | Average number of hours per day the patient experienced itching | 0–24 |
| Itch_Location | Anatomical regions affected by itching, often corresponding to burn sites | None, Trunk, Upper Limbs, Lower Limbs, Combinations |
| QoL_Score | Quality of life score assessed using the ItchyQoL questionnaire, with higher scores indicating worse quality of life due to itching | 26–130 |
| Hospitalization_Duration | Total number of days from admission to discharge | 2–19 |

### 2.4 Statistical Analysis

Statistical analyses were conducted to investigate differences in burn-related outcomes and their relationships with the characteristics of 268 burn patients. Descriptive statistics were computed to summarize the data: means and standard deviations for numerical variables and frequencies for categorical variables (see Table 2).

**Table 2:**
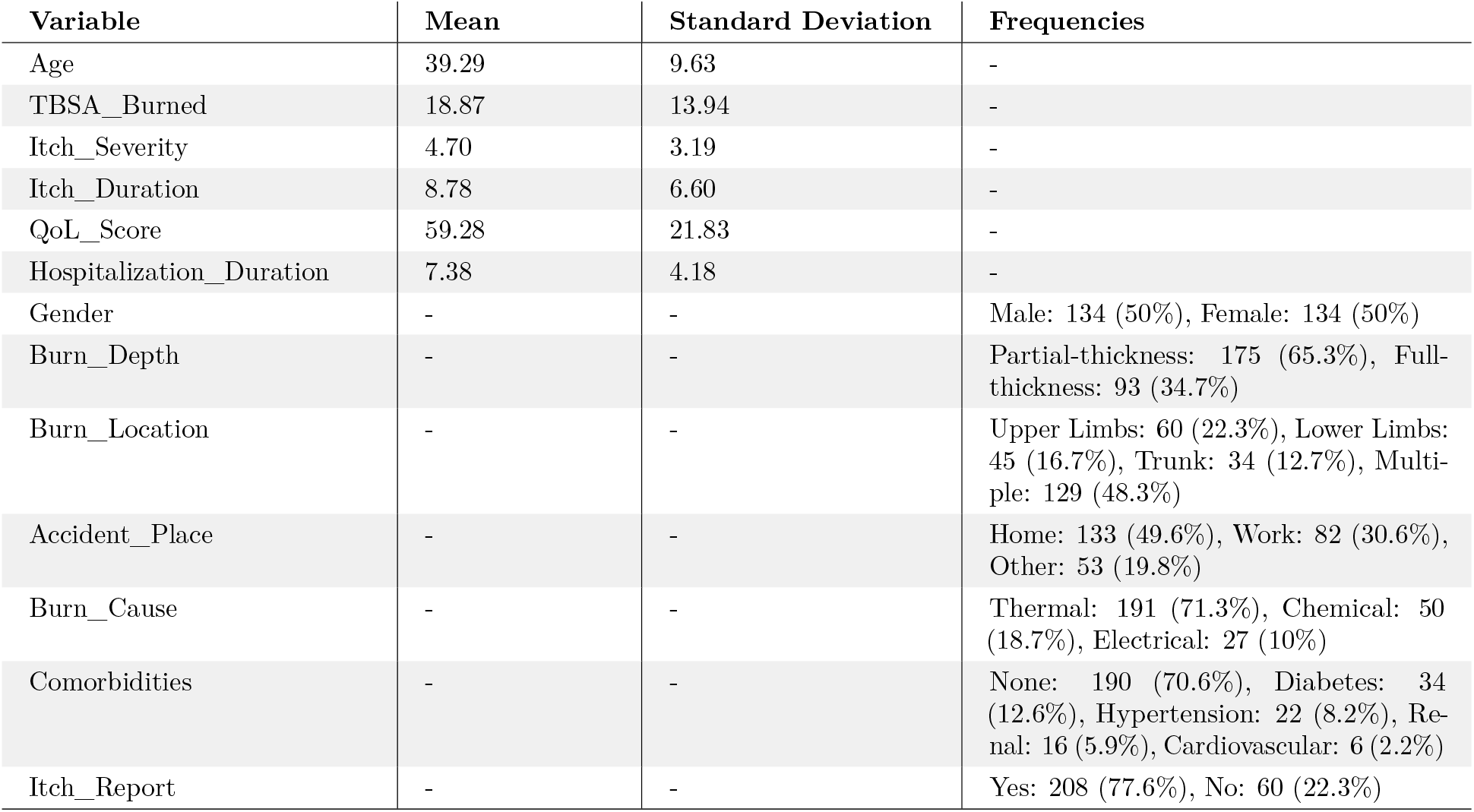
Descriptive Analysis of Dataset Variables. For numerical variables, we provide the mean and standard deviation, and for categorical variables, we report frequencies and range values.

| Variable | Mean | Standard Deviation | Frequencies |
| --- | --- | --- | --- |
| Age | 39.29 | 9.63 | - |
| TBSA_Burned | 18.87 | 13.94 | - |
| Itch_Severity | 4.70 | 3.19 | - |
| Itch_Duration | 8.78 | 6.60 | - |
| QoL_Score | 59.28 | 21.83 | - |
| Hospitalization_Duration | 7.38 | 4.18 | - |
| Gender | - | - | Male: 134 (50%), Female: 134 (50%) |
| Burn_Depth | - | - | Partial-thickness: 175 (65.3%), Full-thickness: 93 (34.7%) |
| Burn_Location | - | - | Upper Limbs: 60 (22.3%), Lower Limbs: 45 (16.7%), Trunk: 34 (12.7%), Multiple: 129 (48.3%) |
| Accident_Place | - | - | Home: 133 (49.6%), Work: 82 (30.6%), Other: 53 (19.8%) |
| Burn_Cause | - | - | Thermal: 191 (71.3%), Chemical: 50 (18.7%), Electrical: 27 (10%) |
| Comorbidities | - | - | None: 190 (70.6%), Diabetes: 34 (12.6%), Hypertension: 22 (8.2%), Renal: 16 (5.9%), Cardiovascular: 6 (2.2%) |
| Itch_Report | - | - | Yes: 208 (77.6%), No: 60 (22.3%) |

To determine the appropriate statistical tests, the normality of numerical variables was assessed using the Kolmogorov-Smirnov and Shapiro-Wilk tests. Variables were considered normally distributed if both tests yielded *p*-values *>* 0.05; otherwise, they were classified as non-normally distributed. Given that key variables, such as TBSA_Burned and Itch_Severity, exhibited non-normal distributions, non-parametric statistical methods were primarily used for analyses.

For comparisons between two independent groups (e.g., Gender: Male vs. Female; Burn_Depth: Partial-thickness vs. Full-thickness), the Mann-Whitney *U* test was employed, as it does not require normality assumptions. For comparisons involving more than two groups (e.g., Burn_Location: Upper Limbs, Lower Limbs, Trunk, or combinations; Burn_Cause: Thermal, Electrical, Chemical), the Kruskal-Wallis *H* test was applied, followed by post-hoc pairwise comparisons when significant differences were detected.

To better understand the relationship between various variables and the quality of life (QoL_Score), linear regression models were applied. First, a standard linear regression using scikit-learn (sklearn) was used to model QoL_Score with predictors such as Itch_Severity, Itch_Duration, TBSA_Burned, and Burn_Depth. This provided a basic linear model to assess the impact of these predictors. In addition, Ordinary Least Squares (OLS) regression was conducted for the same set of predictors, offering detailed statistical insights. These regression analyses are essential for understanding the combined influence of itching and burn characteristics on QoL, which is a primary objective of the study.

Furthermore, correlations between numerical variables—such as QoL_Score and Age, TBSA_Burned, Itch_Severity, Itch_Duration, and Hospitalization_Duration, were evaluated using Spearman’s rank correlation coefficient. This method was chosen because it does not assume normality or linearity, making it well-suited for identifying monotonic relationships.

All statistical tests were two-tailed, with a significance threshold of p *<* 0.05. Analyses were conducted using Python (version 3.10), leveraging the SciPy and Pandas libraries for statistical computations and data manipulation. Results are presented in subsequent sections, highlighting key findings related to burn outcomes and patient quality of life.

### 2.5 Ethical Considerations

This study was approved by the Ethics Committee of the Guilan University of Medical Sciences with the code number IR.GUMS.REC.1401.323. All Participants provided written informed consent, and all procedures followed the principles of the Declaration of Helsinki and its subsequent amendments.

## 3 Results

### 3.1 Normality Assessment

The normality of numerical variables was evaluated using both the Kolmogorov-Smirnov (KS) and Shapiro-Wilk (SW) tests, with the results summarized in Table 3. Age followed a normal distribution (KS test *p* = 0.7729, SW test *p* = 0.1548). However, the other numerical variables (TBSA_Burned, Itch_Severity, Itch_Duration, QoL_Score, Hospitalization_Duration) showed significant deviations from normality (KS test *p <* 0.001, SW test *p <* 0.001). Therefore, non-parametric tests were used for these variables in the subsequent analyses.

**Table 3:** Normality Test Results for Numerical Variables. The normality of the numerical variables was assessed using the Kolmogorov-Smirnov (KS) and Shapiro-Wilk (SW) tests.

| Variable | KS p-value | SW p-value | Normality Interpretation |
| --- | --- | --- | --- |
| Age | 0.773 | 0.155 | Normal ($p > 0.05$) |
| TBSA_Burned | <0.001 | <0.001 | Not Normal ($p \leq 0.05$) |
| Itch_Severity | <0.001 | <0.001 | Not Normal ($p \leq 0.05$) |
| Itch_Duration | <0.001 | <0.001 | Not Normal ($p \leq 0.05$) |
| QoL_Score | <0.001 | <0.001 | Not Normal ($p \leq 0.05$) |
| Hospitalization_Duration | <0.001 | <0.001 | Not Normal ($p \leq 0.05$) |

### 3.2 Impact of Patient Characteristics on Clinical Outcomes

To assess whether numerical outcomes differed significantly across various patient characteristics, we conducted non-parametric statistical tests. Specifically, the Mann-Whitney U test was used for comparisons between two groups, while the Kruskal-Wallis H test was applied for comparisons among more than two groups. These tests were chosen due to the non-normal distribution of the data. The detailed results of these analyses are presented in Tables 4 and 5.

**Table 4:** Results of Mann-Whitney U Tests for Binary Categorical Variables. This table summarizes the results of Mann-Whitney U tests conducted to evaluate differences in continuous outcomes across binary patient characteristics, including gender, burn depth, and the presence of reported itch. The outcomes analyzed were age, total body surface area (TBSA) burned, itch severity, itch duration, quality of life (QoL) score, and hospitalization duration. For each test, the U statistic, p-value, and interpretation regarding statistical significance are reported. A p-value less than 0.05 was considered statistically significant. Associations identified as statistically significant are highlighted in bold within the table.

| Characteristic | Outcome | U Statistic | p-value | Interpretation |
| --- | --- | --- | --- | --- |
| Gender | Age | 8317 | 0.298 | Not Significant |
|  | TBSA_Burned | 8963.5 | 0.982 | Not Significant |
|  | Itch_Severity | 9163.5 | 0.768 | Not Significant |
|  | Itch_Duration | 9606 | 0.319 | Not Significant |
|  | QoL_Score | 9585.5 | 0.339 | Not Significant |
|  | Hospitalization_Duration | 8289.5 | 0.275 | Not Significant |
| Burn_Depth | Age | 7919 | 0.718 | Not Significant |
|  | TBSA_Burned | 8725.5 | 0.33 | Not Significant |
|  | Itch_Severity | 9046 | 0.129 | Not Significant |
|  | Itch_Duration | 8653.5 | 0.39 | Not Significant |
|  | QoL_Score | 9209 | 0.076 | Not Significant |
|  | **Hospitalization_Duration** | **13908.5** | **<0.001** | **Significant** |
| Itch_Report | Age | 6410.5 | 0.748 | Not Significant |
|  | TBSA_Burned | 5563 | 0.169 | Not Significant |
|  | **Itch_Severity** | **0** | **<0.001** | **Significant** |
|  | **Itch_Duration** | **0** | **<0.001** | **Significant** |
|  | **QoL_Score** | **843.5** | **<0.001** | **Significant** |
|  | Hospitalization_Duration | 6343 | 0.845 | Not Significant |

**Table 5:**
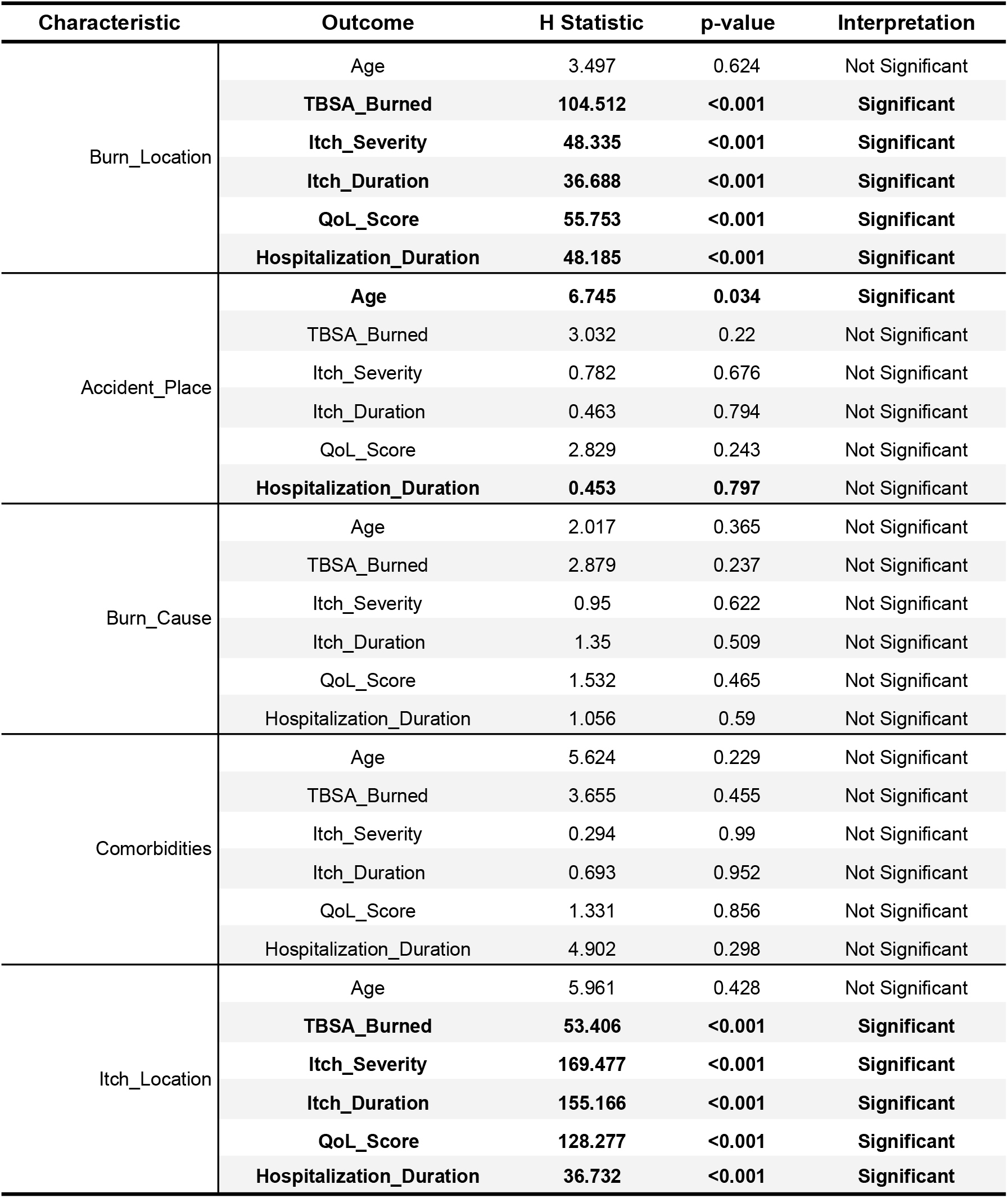
Results of Kruskal-Wallis H Tests for Multi-Categorical Variables. This table presents the results of Kruskal-Wallis H tests conducted to examine differences in continuous outcomes across patient groups defined by multi-categorical variables, such as burn location, accident place, burn cause, comorbidities, and itch location. The outcomes analyzed include age, total body surface area (TBSA) burned, itch severity, itch duration, QoL score, and hospitalization duration. For each comparison, the H statistic, p-value, and the interpretation regarding statistical significance are reported. A p-value of less than 0.05 was considered statistically significant. Associations identified as statistically significant are highlighted in bold within the table.

| Characteristic | Outcome | H Statistic | p-value | Interpretation |
| --- | --- | --- | --- | --- |
| Burn_Location | Age | 3.497 | 0.624 | Not Significant |
|  | **TBSA_Burned** | **104.512** | **<0.001** | **Significant** |
|  | **Itch_Severity** | **48.335** | **<0.001** | **Significant** |
|  | **Itch_Duration** | **36.688** | **<0.001** | **Significant** |
|  | **QoL_Score** | **55.753** | **<0.001** | **Significant** |
|  | **Hospitalization_Duration** | **48.185** | **<0.001** | **Significant** |
| Accident_Place | **Age** | **6.745** | **0.034** | **Significant** |
|  | TBSA_Burned | 3.032 | 0.22 | Not Significant |
|  | Itch_Severity | 0.782 | 0.676 | Not Significant |
|  | Itch_Duration | 0.463 | 0.794 | Not Significant |
|  | QoL_Score | 2.829 | 0.243 | Not Significant |
|  | **Hospitalization_Duration** | **0.453** | **0.797** | Not Significant |
| Burn_Cause | Age | 2.017 | 0.365 | Not Significant |
|  | TBSA_Burned | 2.879 | 0.237 | Not Significant |
|  | Itch_Severity | 0.95 | 0.622 | Not Significant |
|  | Itch_Duration | 1.35 | 0.509 | Not Significant |
|  | QoL_Score | 1.532 | 0.465 | Not Significant |
|  | Hospitalization_Duration | 1.056 | 0.59 | Not Significant |
| Comorbidities | Age | 5.624 | 0.229 | Not Significant |
|  | TBSA_Burned | 3.655 | 0.455 | Not Significant |
|  | Itch_Severity | 0.294 | 0.99 | Not Significant |
|  | Itch_Duration | 0.693 | 0.952 | Not Significant |
|  | QoL_Score | 1.331 | 0.856 | Not Significant |
|  | Hospitalization_Duration | 4.902 | 0.298 | Not Significant |
| Itch_Location | Age | 5.961 | 0.428 | Not Significant |
|  | **TBSA_Burned** | **53.406** | **<0.001** | **Significant** |
|  | **Itch_Severity** | **169.477** | **<0.001** | **Significant** |
|  | **Itch_Duration** | **155.166** | **<0.001** | **Significant** |
|  | **QoL_Score** | **128.277** | **<0.001** | **Significant** |
|  | **Hospitalization_Duration** | **36.732** | **<0.001** | **Significant** |

- **Gender (Male vs. Female)**: No significant differences were observed between males and females for any outcome variables, with *p*-values ranging from 0.275 to 0.912.
- **Burn Depth (Partial-Thickness vs. Full-Thickness)**: Patients with full-thickness burns had significantly longer hospital stays (*p <* 0.001). No significant differences were found for Age, TBSA_Burned, Itch_Severity, or QoL_Score (*p >* 0.05), although a near-significant trend for QoL_Score (*p* = 0.076) suggests a potential reduction in quality of life in this group.
- **Itch Report (Yes vs. No)**: Patients reporting itch had significantly higher Itch_Severity, longer Itch_Duration, and lower QoL_Score (all *p <* 0.001). The U statistic for Itch_Severity and Itch_Duration was 0.0, indicating complete separation between those reporting itch and those not.
- **Burn Location**: Burn location was significantly associated with TBSA_Burned (*p <* 0.001), Itch_Severity (*p* = 0.0054), QoL_Score (*p* = 0.0136), and Hospitalization_Duration (*p <* 0.001). The association with Itch_Duration approached significance (*p* = 0.053), suggesting a potential relationship that might become clearer with a larger sample.
- **Accident Place**: A significant difference in Age was found across accident places (*p* = 0.0344), but no significant differences were observed for TBSA_Burned, Itch_Severity, or QoL_Score (*p >* 0.05).
- **Burn Cause**: No significant differences were observed across different causes of burns, with *p*-values ranging from 0.239 to 0.982.
- **Comorbidities**: No significant differences were found across comorbidity categories for any of the outcomes, with *p*-values between 0.082 and 0.794.
- **Itch Location**: Itch location was significantly associated with TBSA_Burned, Itch_Severity, Itch_Duration, and QoL_Score (all *p <* 0.001), suggesting that the anatomical site of itch had a major impact on burn severity and patient experience.

### 3.3 Correlations with Quality of Life Score

Our regression model demonstrates that the predictors, *Itch Severity* has the most substantial impact (*β* = 3.447), followed by *Burn Depth* (*β* = 1.115), *Itch Duration* (*β* = 0.824), and *Total Body Surface Area (TBSA) Burned* (*β* = 0.486) on QoL. These findings highlight the significant role of itch-related factors in determining post-burn QoL, emphasizing the clinical importance of effective itch management. To further interpret these results, we present a **heatmap** (Figure 1) illustrating the correlations among these variables, confirming the strong association between itch-related measures and QoL. Additionally, **four scatter plots** (Figure 2) visualize the relationships between QoL and each predictor.

**Figure 1:**
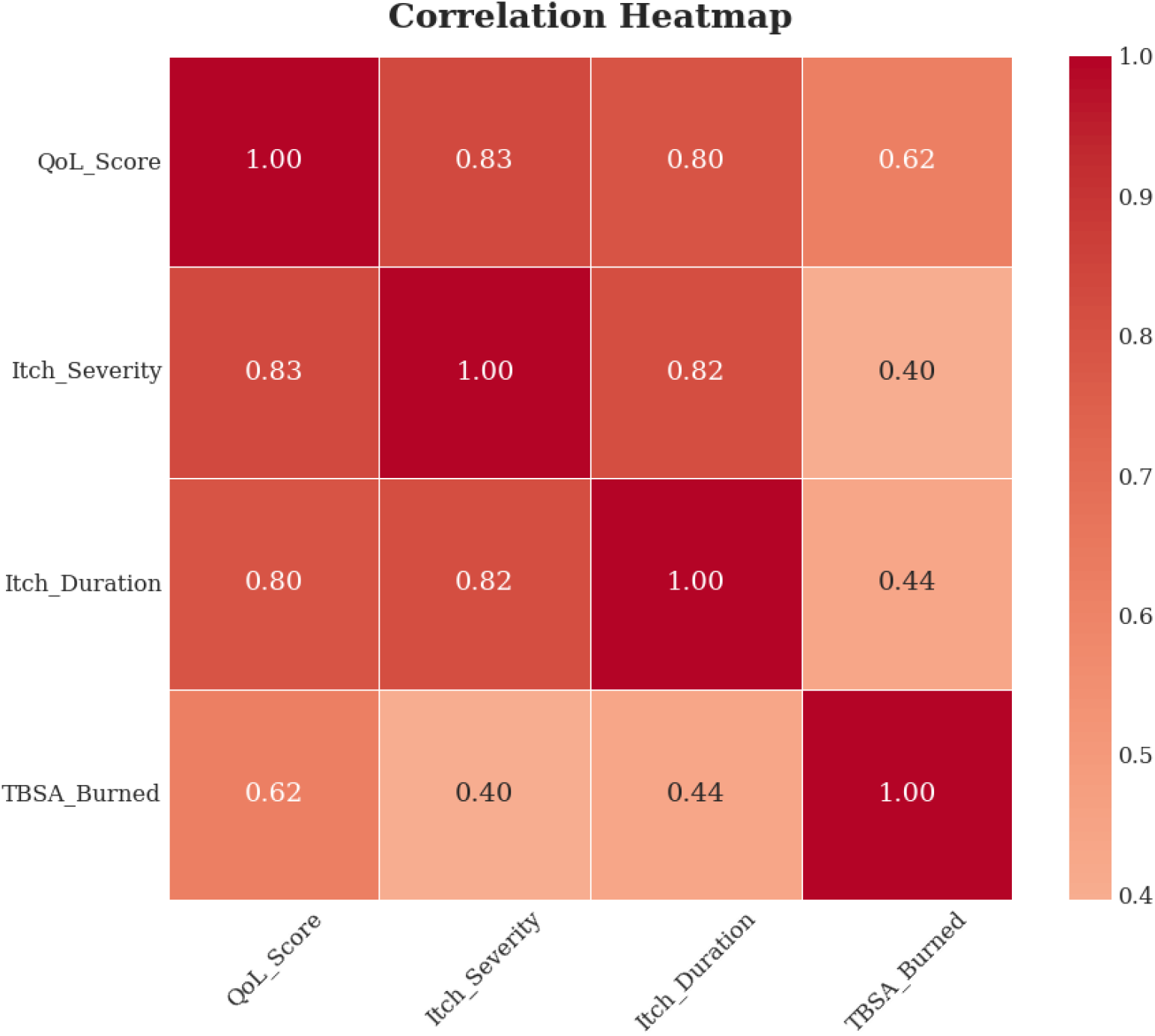
Correlation heatmap of selected variables. This figure illustrates strong positive correlations are observed between QoL_Score, Itch_Severity, and Itch_Duration, while TBSA_Burned shows moderate correlations with the other variables.

**Figure 2:**
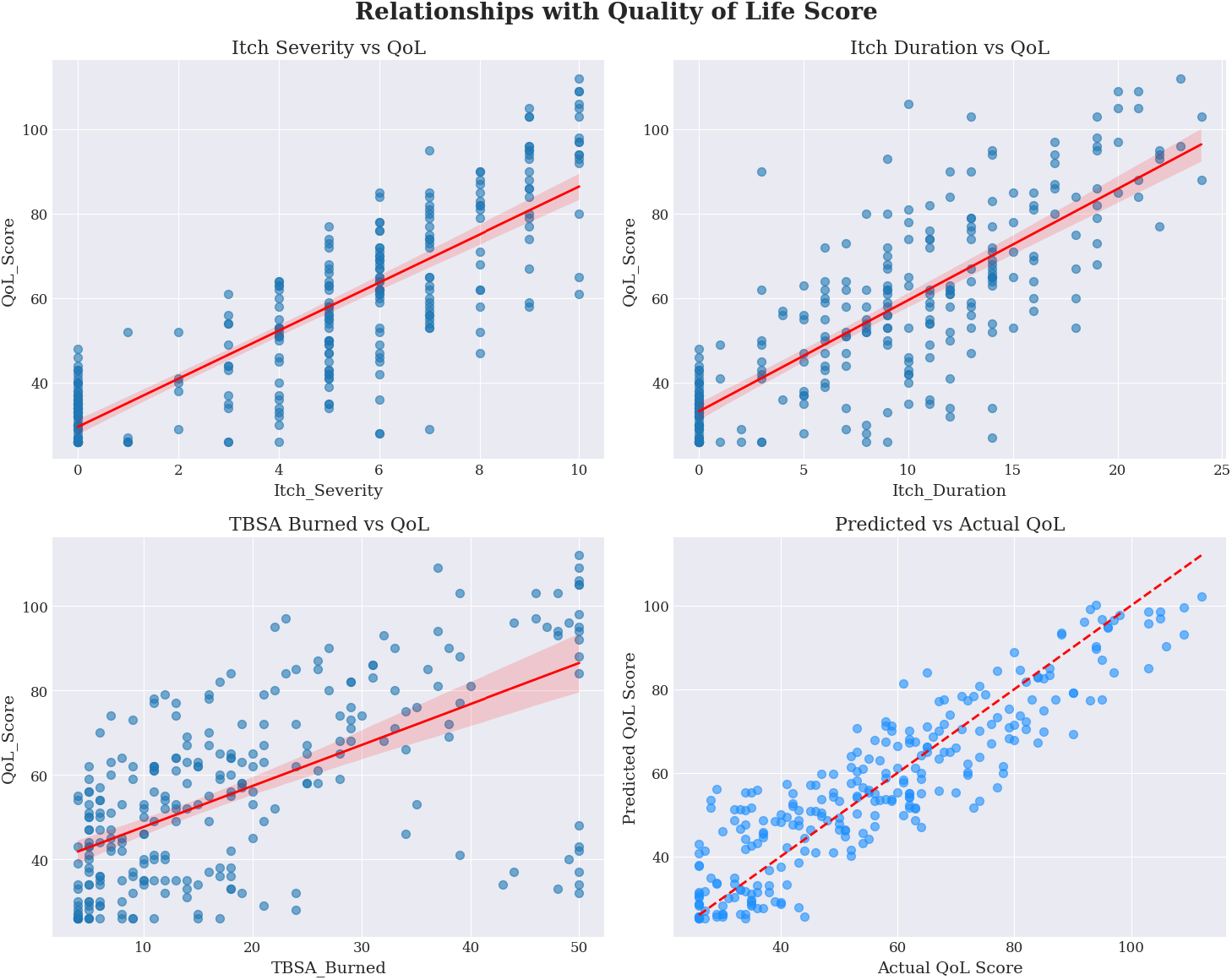
Scatter plots with regression lines. The plots illustrate the relationships between QoL Score and Itch_Severity, Itch_Duration, and TBSA_Burned, alongside the predicted versus actual QoL scores. A positive association is observed in each predictor, with Itch-related variables showing stronger trends compared to TBSA_Burned. The predicted vs. actual plot demonstrates the model’s predictive performance.

Spearman’s rank correlation coefficients were also used to examine the relationships between quality of life and other continuous variables, as shown in Table 6. Significant positive correlations were observed between quality of life and itch severity (*ρ* = 0.805, *p <* 0.001), itch duration (*ρ* = 0.750, *p <* 0.001), and the percentage of body surface area burned (*ρ* = 0.620, *p <* 0.001). These results suggest that more severe itching, longer duration of itch, and greater burn severity are linked to a poorer quality of life. On the other hand, no significant correlation was found between quality of life and patient age (*ρ* = 0.011, *p* = 0.856).

**Table 6:** Spearman’s Rank Correlations Between Quality of Life and Clinical Variables. This table presents the Spearman’s rank correlation coefficients (*ρ*) and associated *p*-values for the relationships between quality of life and various clinical variables, including itch severity, itch duration, burn severity (percentage of body surface area burned), and hospitalization duration.

| Variable | Correlation Coefficient ($\rho$) | p-value |
| --- | --- | --- |
| Age | 0.011 | 0.856 |
| TBSA_Burned | 0.620 | <0.001 |
| Itch_Severity | 0.805 | <0.001 |
| Itch_Duration | 0.750 | <0.001 |
| Hospitalization_Duration | 0.4183 | 0.007 |

In summary, our findings that both the anatomical site of injury and the location of itch significantly affect multiple clinical outcomes—and that itch severity and duration are strongly linked to quality of life—underscore the need to incorporate these considerations into burn-care protocols to optimize patient recovery.

## 4 Conclusion

This study, conducted at Rasht Velayat Hospital, Iran, in 2021, analyzed the effect of acute itchiness during hospitalization on the adult burn patient’s quality of life. Out of the 268 patient population, the itching group experienced greater severity of symptoms, longer time to resolution of discomfort, and poor quality of life than the no-itch patient group. Itch severity was the best predictor of poor quality of life, followed by duration of itch, depth of the burn, and affected percent of the body surface area, according to the regression analysis.

However, the cross-sectional nature of this study limits the ability to capture how itching may evolve over time. Besides, the use of self-report questionnaires makes the study vulnerable to response bias. Since the study was conducted in one hospital, findings may not be extended to large populations of individuals. Longitudinal studies, which would outline the process of post-burn itching, the biological, physiological, and psychological processes of post-burn itching, and develop more effective treatments, particularly the impact of variables such as stress, could be enhanced in future studies.

## Data Availability

All data produced are available online at https://github.com/ramyarfarzan/Impact-of-Acute-In-Hospital-Post-Burn-Pruritus-on-Quality-of-Life-A-Cross-Sectional-Study

## 7 Acknowledgments

The authors are very grateful to the volunteers who participated in this study, the Guilan University of Medical Sciences, Rasht, Iran, and the Velayat Sub-Specialty Burn and Plastic Surgery Center.

## References

[1] Nader Aghakhani, Hamid Sharif Nia, Mohammad Ali Soleimani, Nasim Bahrami, Narges Rahbar, Yadegar Fattahi, and Zahra Beheshti. Prevalence burn injuries and risk factors in persons older the 15 years in urmia burn center in iran. Caspian journal of internal medicine, 2(2):240, 2011.

[2] Ali Bazzi, Mohammad Javad Ghazanfari, Masoumeh Norouzi, Mohammadreza Mobayen, Fateme Jafaraghaee, Amir Emami Zeydi, Joseph Osuji, and Samad Karkhah. Adherence to referral criteria for burn patients; a systematic review. Archives of Academic Emergency Medicine, 10(1):e43, 2022.

[3] P Lynn Bell and Vincent Gabriel. Evidence based review for the treatment of post-burn pruritus. Journal of burn care & research, 30(1):55–61, 2009.

[4] Daniel C Butler, Timothy Berger, Sarina Elmariah, Brian Kim, Sarah Chisolm, Shawn G Kwatra, Nicholas Mollanazar, and Gil Yosipovitch. Chronic pruritus: a review. Jama, 331(24):2114–2124, 2024.

[5] Gretchen J Carrougher, Erin M Martinez, Kara S McMullen, James A Fauerbach, Radha K Holavanahalli, David N Herndon, Shelley A Wiechman, Loren H Engrav, and Nicole S Gibran. Pruritus in adult burn survivors: postburn prevalence and risk factors associated with increased intensity. Journal of Burn Care & Research, 34(1):94–101, 2013.

[6] Bo Young Chung, Han Bi Kim, Min Je Jung, Seok Young Kang, In-Suk Kwak, Chun Wook Park, and Hye One Kim. Post-burn pruritus. International journal of molecular sciences, 21(11):3880, 2020.

[7] Nisha S Desai, Gabriele B Poindexter, Yvette Miller Monthrope, Sandra E Bendeck, Robert A Swerlick, and Suephy C Chen. A pilot quality-of-life instrument for pruritus. Journal of the American Academy of Dermatology, 59(2):234–244, 2008.

[8] J Nicolaas Dijkshoorn, Juanita A Haagsma, Cornelis H van der Vlies, M Jenda Hop, Margriet E van Baar, and Inge Spronk. Assessing health-related quality of life of adult patients with intermediate burns: The added value of an itching and cognition item for the eq-5d: A retrospective observational study. European Burn Journal, 3(2):264–277, 2022.

[9] Food, Drug Administration, et al. Evaluating inclusion and exclusion criteria in clinical trials. In Workshop report. The National Press Club, Washington DC, 2018.

[10] Emilie Fowler and Gil Yosipovitch. Post-burn pruritus and its management—current and new avenues for treatment. Current trauma reports, 5:90–98, 2019.

[11] Karen M Goldstein, Lindsay Chi Yan Kung, Susan Alton Dailey, Aimee Kroll-Desrosiers, Colleen Burke, Megan Shepherd-Banigan, Rebecca Lumsden, Catherine Sims, Julie Schexnayder, Dhara Patel, et al. Strategies for enhancing the representation of women in clinical trials: an evidence map. Systematic Reviews, 13(1):2, 2024.

[12] Kelly AA Kwa, Anouk Pijpe, Zjir M Rashaan, Wim E Tuinebreijer, Roelf S Breederveld, and Nancy EE Van Loey. Course and predictors of pruritus following burns: A multilevel analysis. Acta dermato-venereologica, 98(7), 2018.

[13] Sanaz Masoumi, Marjan Mahdavi-Roshan, Ardalan Majidiniya, Mohammad Ebrahim Ghaffari, Sepideh Pirdastan, Abbas Hajian, and Mohammadreza Mobayen. Effect of probiotic administration in inflammatory responses of thermal burns. Eur. J. Inflamm., 21, June 2023.

[14] Sarah McGarry, Sally Burrows, Tanya Ashoorian, Trisha Pallathil, Katherine Ong, Dale W Edgar, and Fiona Wood. Mental health and itch in burns patients: potential associations. Burns, 42(4):763–768, 2016.

[15] Arun Murari and Kaushal Neelam Singh. Lund and browder chart—modified versus original: a comparative study. Acute and critical care, 34(4):276, 2019.

[16] Bernadette Nedelec and Gretchen J Carrougher. Pain and pruritus postburn injury. Journal of Burn Care & Research, 38(3):142–145, 2017.

[17] Bernadette Nedelec and Leo LaSalle. Postburn itch: A review of the literature. Wounds: a compendium of clinical research and practice, 30(1):E118–E124, 2018.

[18] Grace Obanigba, Jayson W Jay, Steven Wolf, Georgiy Golovko, Juquan Song, Ann Obi, Tsola Efejuku, Dominique Johnson, and Amina El Ayadi. Pre-existing skin diseases as predictors of post-burn pruritus. The American Journal of Surgery, 236:115427, 2024.

[19] Laura KS Parnell, Bernadette Nedelec, Grazyna Rachelska, and Léo LaSalle. Assessment of pruritus characteristics and impact on burn survivors. Journal of burn care & research, 33(3):407–418, 2012.

[20] I Spronks, GJ Bonsel, S Polinder, ME van Baar, MF Janssen, and JA Haagsma. The added value of extending the eq-5d-5l with an itching item for the assessment of health-related quality of life of burn patients: an explorative study. Burns, 47(4):873–879, 2021.

[21] Inge Spronk, Catherine M Legemate, Jan Dokter, Nancy EE Van Loey, Margriet E van Baar, and Suzanne Polinder. Predictors of health-related quality of life after burn injuries: a systematic review. Critical Care, 22:1–13, 2018.

[22] Inge Spronk, Nancy EE Van Loey, Charlie Sewalt, Daan Nieboer, Babette Renneberg, Asgjerd Litleré Moi Caisa Oster, Lotti Orwelius, Margriet E van Baar, Suzanne Polinder, et al. Recovery of health-related quality of life after burn injuries: an individual participant data meta-analysis. PLoS one, 15(1):e0226653, 2020.

[23] Niloofar Tari, Hamidreza Mahmoodi, Seyyed Hamed Hosseini, Kamran Balighi, and Maryam Daneshpazhooh. Structural reliability and validity of persian translation of” quality of life in patients with pruritus (itchyqol)” questionnaire. Dermatology & Cosmetic, 3(2), 2012.

[24] Lincoln M Tracy, Dale W Edgar, Rebecca Schrale, Heather Cleland, Belinda J Gabbe, BRANZ Adult Long-Term Outcomes Pilot Project participating sites, and working party. Predictors of itch and pain in the 12 months following burn injury: results from the burns registry of australia and new zealand (branz) long-term outcomes project. Burns & trauma, 8:tkz004, 2020.

[25] NE Van Loey, HW Hofland, H Hendrickx, J Van de Steenoven, A Boekelaar, and MK Nieuwenhuis. Validation of the burns itch questionnaire. Burns, 42(3):526–534, 2016.

[26] Jie Zhu, Xuanyu Zhao, Alexander A Navarini, and Simon M Mueller. Device-based physical therapies in chronic pruritus: a narrative review. Journal of the American Academy of Dermatology, 2024.

